# PERFORMANCE OF FUJILAM VERSION 1 FOR THE DIAGNOSIS OF TUBERCULOSIS IN CHILDREN UNDER 5 YEARS IN UGANDA – A BRIEF REPORT

**DOI:** 10.1101/2025.10.17.25338231

**Authors:** Mirembe Angella Nanteza, Nelson Mukiza, Freddie Bwanga, Victor Musiime, Grace Paul Kisitu, Nathan Mudrak, Nisreen Khambati, Germine Nakayita, Emmanuel Nasinghe, Prossy Mbekeka, Rudie Desravines, Rutvi Upadhyay, Emily Douglass, Adeodata R. Kekitiinwa, Jerrold J. Ellner, Susan E. Dorman, Adam Penn-Nicholson, Soyeon Kim, David Alland, Padmini Salgame, Rinn Song, Francesca Wanda Basile, Moses Joloba, Else Margreet Bijker, the NOD-pedFEND consortium

## Abstract

**Background:** Diagnosing tuberculosis in children remains challenging due to the paucibacillary nature of the disease, non-specific symptoms, and difficulty collecting samples. New tests are urgently needed. We evaluated the diagnostic accuracy of FujiLAM version 1 urine test in children under 5 years with presumptive tuberculosis in Uganda.

**Methods:** This was a cross-sectional evaluation nested in the NOD-pedFEND diagnostic study. Children under 5 years with signs or symptoms of tuberculosis were recruited from three hospitals in Uganda. All participants underwent comprehensive baseline investigations, including culture and Xpert MTB/RIF Ultra on reference samples and chest X-ray. Culture and Xpert MTB/RIF Ultra results were used to define a microbiological reference standard. FujiLAM version 1 testing was performed on fresh urine at the baseline visit.

**Results:** Seventy-nine children were included in the study. Fifteen participants (19%) were classified as having microbiologically confirmed TB, 44 (56%) as unconfirmed TB, thirteen (16%) as unlikely TB, and seven participants (9%) were unclassifiable. Only one participant (with unlikely TB) had a positive FujiLAM test. Culture and Xpert MTB/RIF Ultra on reference samples were negative in this child, indicating poor diagnostic accuracy with a sensitivity of 0% (95% CI: 0 – 21.8) of FujiLAM version 1 against the microbiological reference standard. *Conclusion:* The poor diagnostic accuracy of FujiLAM version 1 in children under five years of age makes it unsuitable as a diagnostic test in this age group. Further evaluation with the optimized version 2 of this test is needed.

## INTRODUCTION

Diagnosing tuberculosis (TB) in children poses challenges due to its paucibacillary nature and nonspecific symptoms [1]. Additionally, collecting respiratory samples from young children is difficult, and existing tests have limitations in diagnostic accuracy [2]. Thus, improved diagnostic tests for paediatric TB are urgently needed, particularly because the clinical course of TB in children under 5 years can be rapid [3].

Urine-based assays targeting the mycobacterial antigen lipoarabinomannan (LAM) are attractive because they use a non-invasive sample [4]. The Abbott Determine™ TB LAM Ag (Abbott, Chicago, US, previously known as AlereLAM, hereafter Determine TB LAM), has modest sensitivity and specificity and is WHO-endorsed for certain risk groups living with HIV [5]. The Fujifilm SILVAMP TB LAM (FujiLAM; Fujifilm, Tokyo, Japan), is also a urine-based test that detects LAM using high-affinity monoclonal antibodies and silver signal amplification. It has demonstrated moderate sensitivity and specificity, particularly in HIV-positive adult patients [6], but has only been tested in a few paediatric studies [7]. This study assesses FujiLAM’s performance in diagnosing TB in Ugandan children under five years of age.

## MATERIALS AND METHODS

### Study design and population

This prospective cross-sectional diagnostic study was nested within the Novel and Optimized Diagnostics for Pediatric TB in Endemic Countries (NOD-pedFEND) cohort study, which has been described in detail elsewhere [8]. In brief, participant recruitment was conducted at the Baylor Foundation Uganda Center of Excellence (COE) and Mulago National Referral Hospital in Kampala, and the Jinja Regional Referral Hospital in Jinja, all in Uganda. Children under five years of age with presumptive pulmonary TB were prospectively recruited. The sample size was determined by FujiLAM test availability.

### Study procedures

Urine was collected at the baseline visit and tested fresh at the TB laboratory of Makerere University College of Health Sciences using FujiLAM version 1 (lot numbers 20003, 20004 and 21001), as well as Determine TB LAM assays following the manufacturer’s instructions. Because of discrepancies detected when reviewing the Determine TB LAM results, this test was repeated on thawed urine specimens, which were previously frozen at −80 degrees, that were available in 74 children. The final Determine TB LAM result was defined as follows: ‘positive’ if both the initial and repeated results were positive, ‘negative’ if both were negative. All children underwent a thorough clinical assessment, tuberculin skin test, TB contact exposure assessment, chest X-ray, and microbiological reference tests, including Xpert MTB/RIF Ultra (hereafter Xpert Ultra), Mycobacteria growth indicator tube (MGIT), and Löwenstein–Jensen medium (LJ) culture on two nasopharyngeal aspirates, one gastric aspirate, and Xpert Ultra on one stool. They were followed up at 2 weeks, 2 months and 6 months (or end of TB treatment). TB clinical case definitions were assigned based on international consensus definitions [9]. In summary, children were classified as “confirmed TB” if the microbiological reference standard was positive (i.e. at least one mycobacterial culture and/or Xpert Ultra positive on a reference sample), as “unconfirmed TB” in the case of clinical, radiological, and/or immunologic features suggestive of TB but negative microbiological tests, and “unlikely TB” if criteria for confirmed and unconfirmed TB were not met. The remaining children were deemed “unclassifiable” due to missing data.

### Data management and analysis

Demographic and clinical data, HIV status, and diagnostic test results were captured using REDCap. Categorical variables were summarized as frequencies and proportions, continuous variables as medians with interquartile ranges (IQRs), and diagnostic accuracy metrics (sensitivity, specificity, negative predictive value, and positive predictive value) were calculated using simple proportions and 95% confidence intervals using the Clopper-Pearson method. Data were analyzed using STATA 14 and SAS (9.4) (Cary, NC).

### Ethical considerations

Ethics approval was obtained from the Makerere University School of Biomedical Sciences Higher Degree Research Ethics Committee, Uganda National Council for Science and Technology, University Ethics and Compliance at Rutgers, the State University of New Jersey, and the Oxford Tropical Research Ethics Committee (OxTREC). Written informed consent from parents/legal guardians was obtained, and the study procedures adhered to the Declaration of Helsinki.

## RESULTS

### Baseline Characteristics

Seventy-nine children under 5 years of age enrolled from August 2021 to August 2023 in the NOD-pedFEND study underwent FujiLAM version 1 testing. Among them, 44/79 (56%) were male. The median age was 16 months (interquartile range, IQR 12, 30). The median weight-for-length Z score was –1.8 (IQR –3.1, –0.3), while the median weight-for-age Z score was –2.3 (IQR –3.9, –0.7). One participant had missing data on nutrition status; among the remaining 78, 32/78 (41%) were severely malnourished (SAM) according to WHO definitions, 11/78 (14%) were moderately malnourished (MAM), and 35/78 (45%) were not malnourished. Twenty-one (27%) study participants were HIV-exposed but uninfected, 12/79 (15%) were living with HIV, and 46/79 (58%) were HIV-negative and unexposed. At enrolment, 38/79 (48%) were hospitalized. Close contact with TB was reported for 24/79 (30%), while 49/79 (62%) had no known contact, and 6/79 (8%) were uncertain. The majority of the enrolled children, 57/79 (72%), was initiated on TB treatment. Tuberculin skin test (TST) results were positive in 22/79 (28%) and negative in 49/79 (62%), and 8/79 (10%) having invalid results or were not tested (**Error! Reference source not found**.).

### Classification of TB diagnosis

Fifteen participants (19%) were classified as having microbiologically confirmed TB, 44 (56%) as unconfirmed TB, thirteen (16%) as unlikely TB, and seven cases (9%) were unclassifiable. None of the participants were diagnosed with miliary TB.

### Sensitivity, specificity, and predictive values of FujiLAM version 1

Among the 79 children tested with FujiLAM, 78 tested negative, and one tested positive. There were no invalid results. The child who tested positive on FujiLAM was HIV negative and presented with cough, severe acute malnutrition and fever. The child also had a positive Determine TB LAM result, but had a normal X-ray, and symptoms did not improve after initiation of TB treatment, and was therefore classified as having unlikely TB. None of the 15 children with confirmed TB had a positive FujiLAM test, resulting in a sensitivity of 0% (95% CI: 0 – 21.8) against the microbiological reference standard. Out of the 64 children that had no positive culture or Xpert Ultra results on the reference samples, 63 were negative by FujiLAM, indicating a specificity of 98.5% (95% CI: 91.7 – 100) against the microbiological reference standard. The positive predictive value was 0% (95% CI: 0 – 97.5), and the negative predictive value was 81% (95% CI: 70.6 – 89.0). Due to the low number of positive results, subgroup analyses could not be performed.

### Comparison of FujiLAM with Determine TB LAM

Definite Determine TB LAM results were available for 56 participants. The sensitivity of Determine TB LAM was higher than FujiLAM in this cohort, with a positive test result in 2/15 (13%) of the confirmed TB cases and in 8/44 (18%) of the unconfirmed TB cases. Determine TB LAM’s specificity was, however, only 37.5% (95% CI: 8.5 – 75.5), with a negative test in 3/13 unlikely TB cases (Table 2).

**Table 1:**
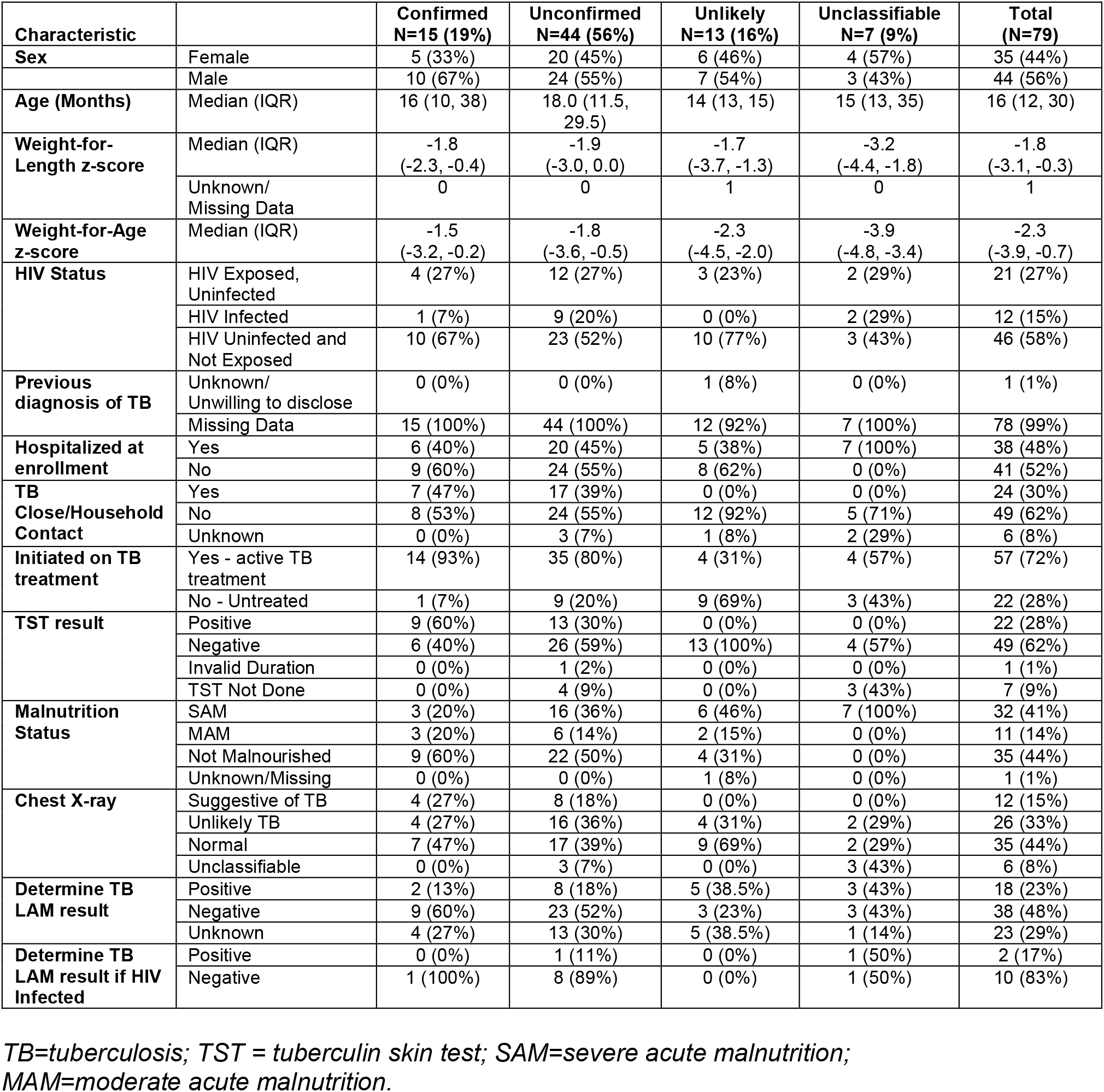
Characteristics of child *participants* with presumed tuberculosis with urine FujiLAM test results (N=79).

**Table 2:** Comparison of FujiLAM version 1 to Determine TB LAM test results in children <5 years of age with different tuberculosis disease categories.

| **Characteristic** |  | **Determine TB LAM result** |  |
| --- | --- | --- | --- |
|  |  | Positive | Negative |
| **NIH Classification** | **Confirmed TB (n=11)** |  |  |
|  | FujiLAM positive | 0 | 0 |
|  | FujiLAM negative | 2 | 9 |
|  | **Unconfirmed TB (n=31)** |  |  |
|  | FujiLAM positive | 0 | 0 |
|  | FujiLAM negative | 8 | 23 |
|  | **Unlikely TB (n=8)** |  |  |
|  | FujiLAM positive | 1 | 0 |
|  | FujiLAM negative | 4 | 3 |
|  | **Unclassifiable (n=6)** |  |  |
|  | FujiLAM positive | 0 | 0 |
|  | FujiLAM negative | 3 | 3 |
| **HIV status** | **Positive (n=10)** |  |  |
|  | FujiLAM positive | 0 | 0 |
|  | FujiLAM negative | 2 | 8 |
|  | **Negative (n=46)** |  |  |
|  | FujiLAM positive | 1 | 0 |
|  | FujiLAM negative | 15 | 30 |

*TB=tuberculosis. Determine TB LAM results were available for 56 participants.*

## DISCUSSION

This study assessed the performance of FujiLAM version 1 in diagnosing TB in children with presumed pulmonary TB under five years of age in Uganda, demonstrating a very low sensitivity (0%) in detecting children with microbiologically confirmed TB. Even though the test’s specificity was high, this challenges its reliability as a diagnostic tool in children under five and risks underdiagnosing TB, leading to missed treatment opportunities. In addition, FujiLAM had no diagnostic yield in HIV-exposed and HIV-positive children, contrary to previous findings, although the number of children was too small to draw definitive conclusions [10, 11].

Prior paediatric studies reported FujiLAM sensitivity ranging from 13% to 42% [12-14]. The lower sensitivity estimates in our study could potentially be influenced by differences in study population, reference standards and clinical settings [4, 7, 10, 11]. Additionally, other studies reported significant lot-to-lot variability of FujiLAM version 1 [15], which has led to the retraction of version 1 from the market by the manufacturer, optimization of the manufacturing process and the launch of version 2 [16].

The study’s strengths included the use of multiple specimens and tests to determine the reference standard (Xpert Ultra, MGIT and LJ mycobacterial cultures) and testing in accredited laboratories. However, its small sample size limited our diagnostic accuracy assessment, and the fact that version 1 of this test will not be brought to the market also limits its wider relevance. In addition, Determine TB LAM assays were repeated, and those discordant were excluded from the comparison of the two LAM assays.

In conclusion, our findings highlight the poor diagnostic performance of FujiLAM version 1 in children under five years of age. Further research with version 2 of this test is needed to refine its potential role in improving paediatric TB diagnosis.

## Data Availability

Data used during this study is available upon reasonable request to the corresponding author.

## Abbreviations

FujiLAM: Fujifilm SILVAMP Tuberculosis LAM
MTB: *Mycobacterium tuberculosis*
HIV: Human Immunodeficiency Virus
NOD: Novel and Optimized Diagnostics
FEND: Feasibility of Novel Diagnostics for TB in Endemic Countries
NOD-pedFEND: the paediatric arm of the FEND study merged with the NOD study
HEPI-SSHU: Health Professional Education Partnership Initiative for Strengthening the Health System and Services in Uganda

## DECLARATIONS

### Ethical Approval and Consent to Participate

Ethics approval was obtained from Makerere University School of Biomedical Sciences Higher Degree Research Ethics Committee, Uganda National Council for Science and Technology, University Ethics and Compliance at Rutgers, The State University of New Jersey, and Oxford Tropical Research Ethics Committee (OxTREC). The study procedures adhered to the standard set by the declaration of Helsinki and Uganda National Council of Science and Technology (UNCST) guidelines for human subject research. To use the secondary data from primary study, a waiver of consent was obtained from School of Medicine Research and Ethics Committee (SOMREC).

### Availability of data and materials

Data used during this study is available upon reasonable request to the corresponding author.

### Authors’ contributions

MAN designed the study, performed data collection and statistical analyses, and wrote the first draft of the manuscript. NM supported data collection and analysis. VM, FB and EB contributed to study conception and results interpretation. GPK, NM, NK, GN, EN, PM, RD, RU, ED, SK and FWB contributed to collection and/or analysis of data. AK, JE, SED, APN, DA, PS, RS and MS were principal investigators of the parent study. All authors read and approved the final manuscript.

#### NOD-pedFEND consortium authors

Gerald Agaba Muzorah, Gideon Ahimbisibwe, Sharley Melissa Aloyo, Sheillah Ansiima, Derek Armstrong, Kiranjot Arora, Sandra Ruth Babirye, Henry Balwa, Kisegerwa Bashir, April Borkman, Eric Bugumirwa, Tatiana Caceres, Rodrigo Calderon, Andrea Cavallini, Ted Cohen, Nandini Dendukuri, Margaretha De Vos, Uzochukwu Egere, Christie Eichberg, Karla Giannina Ali Francia, Steve M. Graham, George Haumba, David Hom, Pitchaya Peach Indravudh, Farag Kakyama Luwambya, Florence Kalawa, Florence Kalembe, Kanyange Angel Moureen, Samuel Kasibante, Nakitto Aisha Kawwoya, Sandra V. Kik, Malik Koire, Kyamulabi Princess Daisy, Yhanella Lagos, Nair Lovaton, John Paul Lubega, Rose Nabatanzi, Agnes Malobela, Ben J. Marais, Frank Matovu, Nicolas Menzies, Francisco Mestanza, Carolina Moron, Rita Makabayi Mugabe, Benedicto Mugabi, Nelson Muhumuza, Levi Mwanga, Rose Nabirye, Allen Nabisere, Stephannie Nabuduwa, Mary Nakagwa, Brenda Sharon Nakalanda, Lydia Nakiyingi, Rose Namaganda, Aminah Nakyazze, Hellen Nassolo, Claire Night, Gloria Ninsiima, Israel Odongo, Laura Olbrich, Megan Palmer, Silvia Perez Romero, Rebecca Post, Kamulegeya Rogers, Morten Ruhwald, H. Simon Schaaf, Ian Schiller, Willy Ssengooba, Sedona Sweeney, Abner Tagoola, Jessica Tagobera, Ann Tufariello, Agnes Turyamubona, César Ugarte-Gil, Luz Villa, Eric Wobudeya, Yingda Xie, Marjorie Yupanqui, Carlos Zamudio.

## Acknowledgements

The authors thank the study participants and their parents/caregivers for making this study possible, the Baylor Foundation Uganda and Jinja Regional Referral Hospital staff for contributing to data collection, and the Mulago Hospital staff for contributing to the laboratory analyses.

